# Bayesian Network Modelling for the Clinical Diagnosis of Alzheimer’s Disease

**DOI:** 10.1101/2023.12.30.23300452

**Authors:** Amber-Rose Bate St Cliere, Norman Fenton

## Abstract

Alzheimer’s disease (AD) is a progressively debilitating disease commonly affecting the elderly. Correct diagnosis is important for patients to access suitable therapies and support that can help improve or manage symptoms of the condition. Reports of misdiagnosis and difficulty diagnosing AD highlight existing clinical challenges. Here we propose a Bayesian network as a preliminary model for a complementary clinical diagnostic tool for dementia due to AD and mild cognitive impairment due to AD. The model structure was built based on medical reasoning patterns which help bridge the gap between clinical professionals and algorithmic decision making. The parameters of the model were specified from a combination of learning from data (using the NACC Uniform Data Set), extracting data from literature, and knowledge-based judgment. The resulting model includes variables laid out in NIA-AA diagnostic criteria and differentiates actual AD cases from formal AD diagnoses. The model is validated against a range of real-world data. Unlike machine-learnt (black box) AI models, this model provides a visible and auditable justification for its predictions and can be used for multiple types of ‘what if analysis’. An easy-to-use web accessible version of the model has been made available.

## 1. Introduction

Alzheimer’s Disease (AD) is a neurodegenerative disease that commonly affects the elderly population. It is characterised by the presence of plaques and neurofibrillary tangles in the brain, primarily composed of proteins called amyloid beta (Aβ) and tau, respectively^1^. The symptoms are progressively debilitating, which impacts the quality of life for affected individuals and their caregivers^2^. Most AD patients develop symptoms later in life (typically at or above 65 years of age) which is referred to as late-onset AD. Early-onset AD occurs in a smaller subset of AD patients who typically develop symptoms in their 30s, 40s and 50s^3^. There are three broad phases of AD:

1. Preclinical AD: asymptomatic but possible biological brain changes^4^.
2. Mild cognitive impairment (MCI) due to AD: mild symptoms, but the affected individual maintains independence in daily life^5^ (referred to hereafter as AD MCI).
3. Dementia due to AD: symptoms interfere with daily living activities^4^ (referred to hereafter as AD dementia).

AD is the leading cause of dementia, a general term for impairment in memory and other cognitive domains that is severe enough to impair daily living activities^6^. There are several other causes of dementia, which are treated and managed differently^7^. Therefore, the correct diagnosis is important. It is estimated that AD dementia affects 11.3% of those aged 65 and above^8^. With an ageing population and age being a major risk factor for AD, prevalence is projected to rise^9^. AD diagnosis enables the patient to access required support as well as drug and non-drug therapies that could potentially stabilise or improve the condition^10^. Despite this, there are shortcomings in the clinical diagnosis of AD. The Alzheimer’s Association 2019 survey found that 90% of primary care physicians believe it is important to diagnose AD MCI but 51% are not fully comfortable diagnosing it^4^, while nearly 40% reported that they were “never” or “only sometimes” comfortable diagnosing AD or another dementia^11^. Moreover, studies estimate that between 12% and 23% of those diagnosed with AD are misdiagnosed^12^. A plausible reason for this is that the symptoms of AD are not specific to one disease and can be caused by numerous conditions. This includes other causes of dementia (such as frontotemporal dementia and dementia with Lewy bodies) and reversible conditions (such as drug abuse and nutrient deficiency)^13^.

The objective of this work is to develop improved AD diagnosis using a causal Bayesian Network (BN) that combines knowledge and data. BNs are a favourable tool for medical diagnosis due to their interpretability^14^. Unlike pure machine learning models, which are essentially black box models^15^, the proposed causal BN model has a structure that is understandable to clinicians and lay people. The primary function of the model is to provide a probability of suspected AD (the probability that an individual has the disease) as well as the probability they will be formally diagnosed with the disease. The distinction between *having the disease* (a hypothesis that is generally never knowable for certain) and *being diagnosed with the disease* is critical both in practice and in the proposed model. These results are calculated through a Bayesian approach which updates the prior probability that an individual has AD when new information is observed. The BN is built using a causal structure based on medical reasoning patterns (referred to as medical idioms) as proposed by Kyrimi *et al*.^14^. The parameters are specified from a combination of learning from data (using the National Alzheimer’s Coordinating Center Uniform Data Set (NACC UDS)), extracting data from the literature, and knowledge-based judgement.

The rest of this paper is structured as follows: in section 2 we review the literature on related AI approaches for AD diagnosis. In section 3 we provide an overview of BNs and describe the methodology used to build the BN. Section 4 describes the model validation and provides results of the BN under example scenarios. Conclusions, discussions, and suggestions for future work are given in section 5.

## 2. Related AI approaches Alzheimer’S Disease

Clinical decision support systems that leverage machine learning have been increasingly developed and deployed^16^. Several machine learning methods have been used to model the progression of AD including neural networks, support vector machines (SVMs), regression and decision trees^17^. These models are primarily data-driven, and the clinician’s interpretation is hindered by the underlying complex algorithmic processes. Moreover, these models do not distinguish causal relationships between variables. Recently developed and advanced AI tools, such as ChatGPT, have demonstrated potential for medical disease diagnosis. Nevertheless, it is crucial to establish proper regulatory guidelines to determine its role in clinical practice, which are currently not fully defined for such Neural Language Processing-based tools^18^. Moreover, the ‘black box’ nature of the ChatGPT model raises concerns regarding its interpretability for healthcare professionals. Using a BN enables us to implement a causal model that follows medical reasoning patterns and thus is an intuitive choice for clinical decision support systems. Hence, we restrict our focus here on reviewing Bayesian approaches for the detection and diagnosis of AD, although they have also been employed specifically for the prediction of biomarkers^19^ and efficacy of treatment for AD^20,21^.

There have been several Bayesian models proposed that were not causal BN models^22^ including naïve Bayes models^23^. Recently García-Gutierrez *et al*. proposed a two-layered model for the diagnosis of AD and Frontotemporal Dementia (FTD)^24^. The first layer executes binary classification using SVMs. The second layer takes the output from the first layer and performs multiclass classification using either evolutionary grammars or BNs. This work has several strengths including addressing another form of dementia that can be misdiagnosed AD. However, the modelling strategy used is non-trivial, where BNs are only a possible component alongside other machine learning models.

A number of previous approaches have relied solely on a BN approach. Out of the BN models for AD diagnosis, two predate the updated AD diagnostic 2011 criteria and do not include various symptom and biomarker variables important for the diagnosis of AD^22,25^. In 2016 Seixas *et al*. proposed a BN model for the diagnosis of AD, dementia, and MCI^26^. It was noted that the model did not include biomarker tests which can be used to support AD diagnosis. Moreover, all-cause MCI was used rather than AD MCI specifically. Alexiou *et al*. proposed a model for the early diagnosis of AD at different stages of the disease^27^. The variables included in the model almost exclusively consisted of biomarkers, some of which would not be used in clinical settings. Guerrero *et al.* developed a BN to be used as a screening tool for cognitive impairment compatible with early stages of AD^28^. The model was based solely on identifying semantic memory impairment. Pillai and Leong proposed a BN for detecting AD where the structure was learnt from data with guidance from experts^29^. Genetic variables that are not used in clinical settings were included while several clinically relevant variables were not included. It is worth noting that Bayesian approaches have been used in the context of medical images for AD diagnosis, such as that proposed by Illan *et al*. and Payares-Garcia *et al.* using MRI brain images^30,31^. However, the work we present in this paper does not involve image processing and classification.

Several of the proposed BNs do not include clinically relevant variables and lack a structure that adheres to causal reasoning patterns (medical idioms). Moreover, all the models reviewed specified a single node for the condition (the diagnosis) whereas the critical distinction between having the condition and a formal diagnosis can account for scenarios where an individual may be misdiagnosed or simply undiagnosed. Where age was included as a variable, the distinction between early-onset AD and late-onset AD was not apparent, thus making these models specific to the diagnosis of late-onset AD. Our proposed BN is an improvement on the state-of-the-art since it is an easy-to-use diagnostic tool that is applicable for both late and early-onset AD, includes all relevant clinical variables, and applies medical idioms to aid AI interpretability.

## 3. Methodology

### 3.1 Requirements Capture

The BN should function as a complementary diagnostic tool to aid the decision making of medical professionals. To comply with clinical practice, the BN for the diagnosis of AD includes variables laid out in validated AD diagnostic criteria. Section 3.1.1 presents an overview of BNs. An outline of diagnostic criteria for AD is provided in Section 3.1.2.

#### 3.1.1 Bayesian networks

A BN is a graphical probabilistic model presented as a directed acyclic graph^32^. The graph comprises nodes (representing variables) that are connected via directed arcs that represent a causal or influential relationship. Building the structure of a BN involves choosing which variables to include and where to position the arcs between nodes, including the arc direction. Figure 1a shows an example three-node BN. A family history of AD has a causal effect on developing the disease. Thus, the direction of the arc is from ‘*Family History’* to ‘*Suspected AD’*. This makes ‘*Family History’* the ‘parent’ node of ‘*Suspected AD’*, and ‘*Suspected AD*’ the ‘child’ node of ‘*Family History*’. Likewise, AD causes the symptom of memory impairment which is reflected in the structure of the three-node example. Each node has a Node Probability Table (NPT) which represents the conditional probability distribution of a node given its parents (Figure 1b).

**Figure 1.**
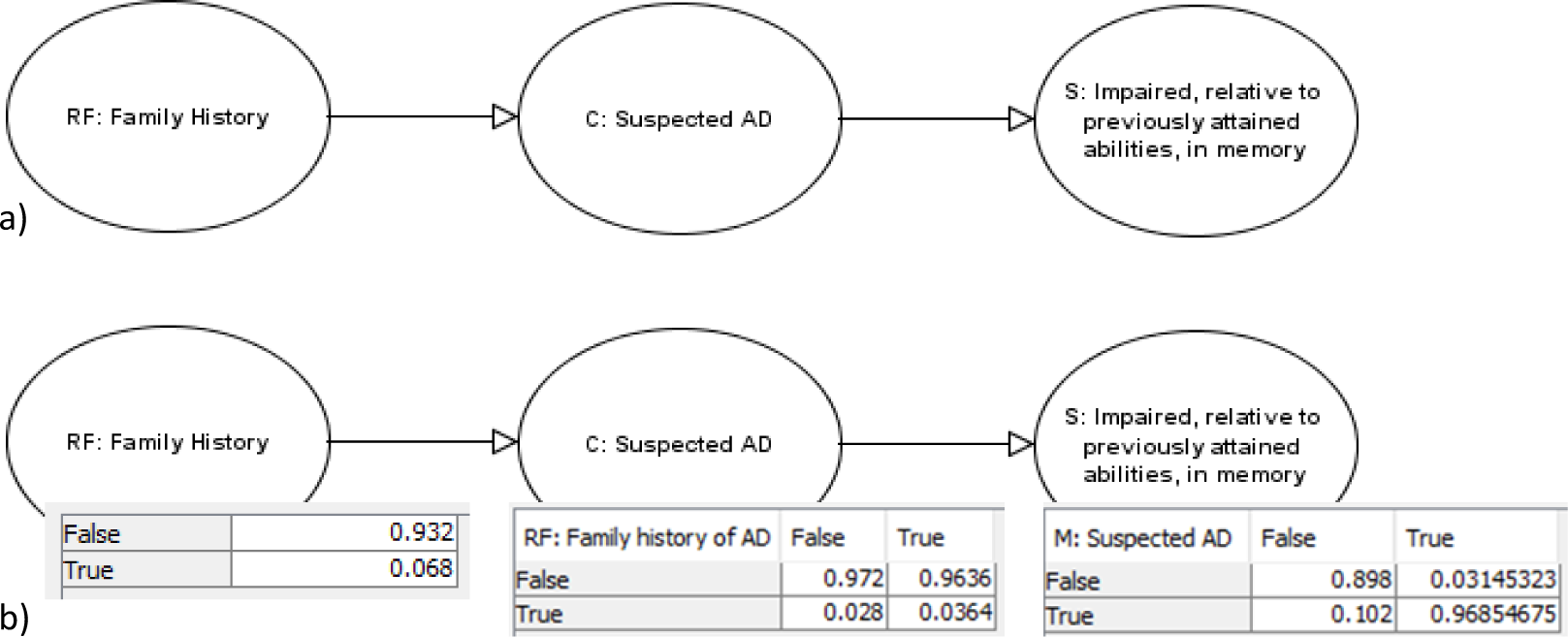
(a) A three-node Bayesian network. (b) A three-node Bayesian network with Node Probability Tables shown.

As an example, the NPT of Suspected AD is:

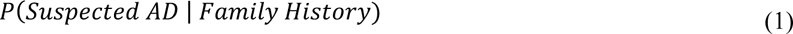

where *Suspected AD* and *Family History* both have two states, True or False.

To parametrise the BN, values for all the NPTs must be provided. This enables Bayesian probabilistic reasoning whereby we update our prior belief of an uncertain hypothesis when new information or evidence is observed. The prior belief is referred to as the *prior probability* and the updated belief is referred to as the *posterior probability*. For example, to calculate the posterior probability:

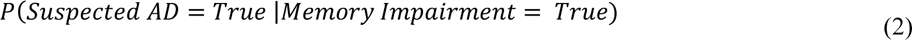

the BN would perform the following calculation:

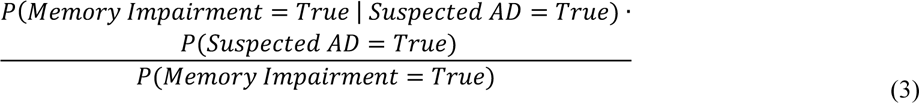

Where *P(Memory Impairment = True)* is the new evidence observed and *P(Suspected AD = True)* is the prior probability.

An advantage of BNs for use in medical settings is that once evidence is observed (the state of a node is entered), the probabilities for the remaining unobserved variables are updated. This occurs through forward and backward reasoning. The former process follows the direction of the arc, and the latter follows the counter direction. A BN will produce an updated prediction no matter how few, or many variables are observed. This means that we can use the model for early diagnosis (e.g., based only on a patient’s basic physical/demographic attributes) and can update the probability of the diagnosis after we observe symptoms and test results. Another advantage is the ability to model interventions, such as estimating the probability a patient will exhibit depressive symptoms if they take antidepressant medication.

#### 3.1.2 Diagnostic Criteria for Alzheimer’s Disease

The BN for AD diagnosis should contain variables outlined in validated clinical diagnostic criteria. Several associations have proposed AD diagnostic criteria, including the Diagnostic and Statistical Manual of Mental Disorders^33^, the International Working Group^34^ and the National Institute for Aging-Alzheimer’s Association (NIA-AA). The National Institute for Health and Care Excellence (NICE) guidelines recommend the use of NIA-AA criteria^7^.

The National Institute of Neurological and Communicative Disorders and Stroke (NINCDS) and the Alzheimer’s Disease and Related Disorders Association (ARDA) outlined the diagnostic criteria for AD dementia in 1984. These criteria were revised and expanded to recognise the two stages preceding AD dementia (preclinical AD and AD MCI) by the NIA-AA in 2011. The NIA-AA presents the following articles for AD: towards defining the preclinical stages of AD^35^, the diagnosis of MCI due to AD^5^, and the diagnosis of dementia due to AD^36^. It should be noted that there is a distinction between diagnostic criteria for clinical versus research purposes (for example in clinical trials). The proposed criteria for defining the preclinical stages of AD are intended solely for research purposes. Thus, the BN for clinical diagnosis of AD will only consider AD MCI and AD dementia.

The criteria for definitive AD according to the NINCDS-ARDA 1984 guidelines are: “the clinical criteria for probable AD and histopathologic evidence obtained from biopsy or autopsy”^37^. The 2011 NIA-AA criteria do not offer an update for the diagnosis of definitive AD, though they do expand the guidelines for probable AD with pathological evidence. A simplification of the NIA-AA clinical criteria for probable AD dementia can be summarised as fulfilling the following:

1. Meeting the criteria for dementia.
2. Gradual onset of symptoms.
3. History of worsening cognition.
4. Initial and most prominent cognitive impairment in either memory, language, visuospatial function, or executive function. Impairment in at least two cognitive domains should be present.

Evidence of a pathogenic mutation in amyloid precursor protein (*APP*), presenilin 1 (*PSEN1*), or presenilin 2 (*PSEN2*) genes increases the level of certainty for probable AD dementia. Furthermore, if an individual meets the core clinical criteria for probable AD dementia, positive biomarker tests can increase the certainty that the AD pathology is the basis of dementia.

The NIA-AA criteria also specify that probable AD dementia should not be diagnosed when there is evidence for another neurological disease or non-neurological condition that can have a substantial effect on cognition. In this case, an individual may instead meet the criteria for possible AD dementia. The criteria for possible AD dementia also include those who meet the criteria for probable AD dementia except the onset of symptoms is sudden rather than gradual. The key difference between AD dementia and AD MCI is that the criteria for AD MCI is met when the individual maintains independence of function in daily life. See ^5,36^ for a more detailed description of criteria.

### 3.2 Design and Implementation

The following section presents the main design elements of the BN and how they were implemented. We describe how the BN was structured according to medical idioms in section 3.2.1. Section 3.2.2 provides an overview of how the parameters were specified, including use of the National Alzheimer’s Coordinating Center (NACC) dataset. The model was built using agena.ai software (www.agena.ai).

#### 3.2.1 Structuring the Bayesian Network

Work by Kyrimi e*t al*. proposes a set of medical idioms that were used to inform the BN model structure^14^. Variables are classified based on their role in clinical reasoning as follows: Condition (C), Manifestations (M), Risk Factors (RF), Treatment (T), Comorbidities (CC) and Complications (Cm). Manifestations are observable consequences of a condition and can be divided into Symptoms (Sy), Signs (Si) and Medical Tests (Mt)^14^. Once variables are classified and the relationships are determined, the respective medical idioms can be applied to build a structure like that shown in Figure 2.

**Figure 2.**
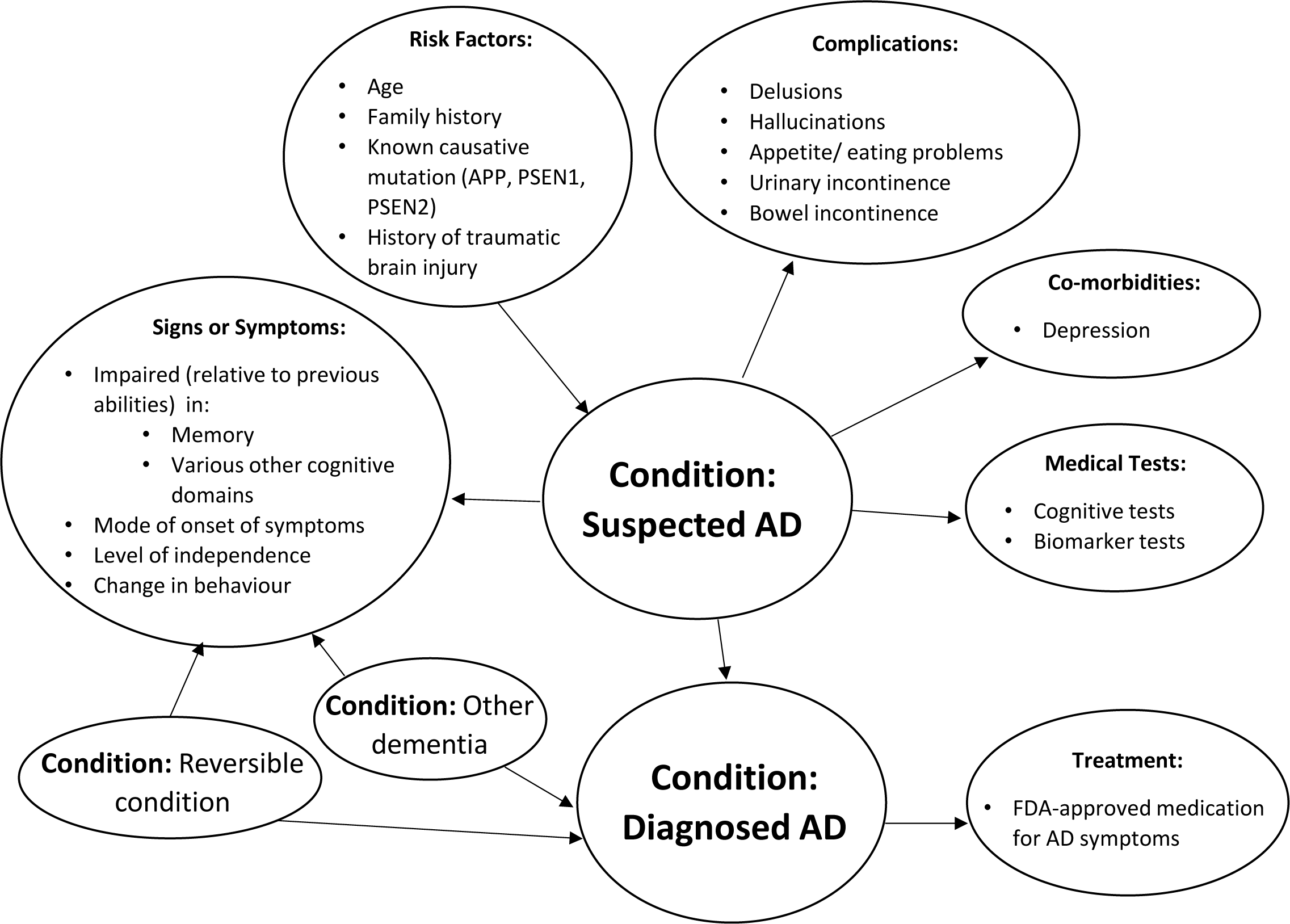
Simplified model schematic with the main variables included in the Bayesian network. Note that this is not the full model structure.

##### Conditions, Signs and Symptoms

The main condition is AD, and there is a crucial distinction between ‘*C: Suspected AD’* and ‘*C: Diagnosed AD’*, the former refers to having the disease and the latter refers to being formally diagnosed. Note that if a person who genuinely has AD is not subject to any assessment or tests then, of course, they will not be formally classified as diagnosed with AD; this is a classic problem pertinent to all AI models that rely only on data which assumes the only people classified as diagnosed/confirmed cases of the disease are those who have the disease. To account for this the node ‘*Formal Testing’* was included as a parent to ‘*C: Diagnosed AD’,* when the former is observed as False, the latter will also be False. The manifestation idiom models the uncertain causal relationship between a condition and the variables for signs, symptoms, and medical tests. Thus ‘*C: Suspected AD’* is a parent node to the signs, symptoms, and medical tests nodes. The distinction between signs and symptoms is difficult to define in the context of AD. The NICE guidelines recommend getting an account of the symptoms from the person with suspected dementia as well as someone who knows the person well, referred to as an informant^7^. While the manifestations of AD, including memory impairment, can be thought of as symptom (a subjective feeling which is only apparent to the patient)^14^ this may be observed by an informant and/or the clinician as a sign. For simplification, Signs and Symptoms will be consolidated to Signs or Symptoms (S). As outlined by the NIA-AA criteria, the hallmark symptom of AD is amnestic presentation, referring to memory impairment. Additional symptoms include impairment in language, visuospatial function, executive function, orientation, and attention or concentration^5,36,38^. AD patients may also exhibit changes in personality or behaviour^39^. Another sign from the NIA-AA criteria is that the mode of onset of symptoms is gradual as opposed to sudden. The distinction between AD MCI and AD dementia relies on level of independence^5,36^.

AD diagnostic criteria also specify that the diagnosis of AD is impacted by evidence of a concurrent, neurological, or non-neurological condition that can have a substantial effect on cognition^36^. This includes other conditions that cause dementia as well as reversible conditions that can be attributed to as the cause of cognitive impairment. Therefore, the variables ‘*C: Other dementia’* and ‘*C: Reversible condition’* are included in the BN as parent nodes to both the signs or symptoms nodes and to ‘*C: Diagnosed AD’.* For simplicity we have separated other causes of cognitive impairment into dementia conditions and reversible conditions that cause cognitive impairment. The distinction between the two categories relies on whether the condition is reversible. For example, delirium and side effects of medication are reversible conditions whereas vascular dementia and AD are not. For details on all conditions included in the two categories see Appendix A Table 1.

**Table 1.** Full names and explanations for certain variables included in the BN.

| Variable/Node Name (in the Model) | Variable Description |
| --- | --- |
| C: Other Dementia | Diagnosis of/ strong evidence for either a dementia causing disease other than AD, or of a progressive neurological disease that can affect cognition but is not necessarily considered dementia. This includes:<br>1. Frontotemporal dementia (FTD) / Frontotemporal lobar degeneration (FTLD), including diagnoses of Primary Progressive Aphasia (PPA), behavioural variant FTD, FTLD with motor neuron disease or FTLD not otherwise specified<br>2. Lewy bodies dementia<br>3. Vascular dementia<br>4. Prion disease (CJD and others)<br>5. Huntingdon's disease<br>6. Parkinson's disease<br>7. Posterior cortical atrophy (PCA)<br>8. Progressive supranuclear palsy (PSP)<br>9. Corticobasal syndrome (CBS)<br>10. Cerebrovascular disease, defined by the presence of either a single strategic infarct, multiple infarcts, or extensive white matter hyperintensity<br>11. Multiple Systems Atrophy (MSA)<br>This variable is not mutually exclusively with other condition variables. |
| C: Reversible Condition | Diagnosis of a reversible condition that can affect cognition. This includes:<br>1. Post-traumatic stress-disorder (PTSD)<br>2. Alcohol abuse<br>3. Other substance abuse<br>4. B12 deficiency |
|  | 5. Sleep disorder (sleep apnea, RBD, insomnia)<br>6. Side effects from medication<br>7. Delirium<br>8. Normal pressure hydrocephalus<br>9. CNS neoplasm<br>This variable is not mutually exclusive with other condition variables. |
| S: Memory/Language/Visuospatial Function/Executive Function/Orientation/Attention or Concentration/Other Cognitive Domain Impairment | Impaired, relative to previously attained abilities, in Memory/Language/Visuospatial Function/Executive Function (judgment, planning or problem solving) /Orientation/Attention or Concentration/Other Cognitive Domains. |
| S: Change in Behavior or Personality | Subject currently manifests meaningful change in behavior including any of the following: apathy and withdrawal, irritability, disinhibition, agitation. Or subject exhibits a change in personality. |
| T: Current use of AD medication | Current use of FDA-approved medication for AD symptoms. |

##### Medical Tests

Medical tests can be divided into mental cognitive status tests and biomarker tests. According to the Alzheimer’s Association, The Mini-Mental State Exam (MMSE) is commonly used for AD diagnosis^40^. The Clinical Dementia Rating (CDR® Dementia Staging Instrument) and the Montreal Cognitive Assessment (MoCA) were also included. Regarding biomarker tests, the NIA-AA state they are optional tools to be used where available and when the clinician considers them appropriate.^36^ Since these criteria were published in 2011, there has been several reports on the advantage of using biomarker tests for the clinical diagnosis of AD and are more frequently used in clinical settings^7,41,42^. For example, the amyloid imaging task force report outlines appropriate clinical use for Positron Emission Tomography (PET) amyloid imaging. They further state that incorporating this into clinical decision making for AD may help delineate AD MCI from AD dementia^43^. Biomarker tests include: low cerebrospinal fluid (CSF) Aβ42, PET amyloid imaging, elevated CSF tau or phosphorylated tau, decreased fluorodeoxyglucose (FDG) uptake on PET, and disproportionate hippocampal atrophy on structural MRI^36^. The node ‘*C: Suspected AD’* is causal of the medical test results and thus is a parent node to all medical test nodes. The cognitive tests are also child nodes to ‘*C: Other dementia*’, ‘*C: Reversible condition’* and ‘*65 and above’* (the latter due to age-associated memory impairment^44^). Biomarker test results are also child nodes to ‘*65 and above’*, as the probability of exhibiting biomarker signs of AD increases with age, even in cognitively normal patients^45^.

##### Comorbidities, Complications and Dementia Severity

Several comorbidities of AD have been suggested, including cardiovascular disease and diabetes^46^. Though it is unclear as to whether these are risk factors or comorbidities. Depression is commonly observed in AD and other dementia patients and was included as a comorbidity in the BN as a child node to ‘*C: Suspected AD*’ and ‘*C: Other dementia’*. It is a parent node to ‘*Depression or dysphoria severity’* which is a measure of the symptoms the patient experiences. An analysis on the National Health and Aging Study found that engaging in a favourite activity is associated with lower levels of depression and greater functional independence in dementia patients^47^. Other studies have found activity engagement to be associated with positive mood in people with dementia, with the type of activity being less important than activity engagement itself^48^. Therefore, the node ‘*Dropped many activities and interests’* is included as a parent to the ‘*Depression or dysphoria severity’* node.

AD dementia has several unfavourable consequences (termed Complications: Cm in the BN) including problems with eating and appetite, urinary incontinence, bowel incontinence, delusions, and hallucinations^39,49^. Complications are intrinsically linked to the severity of dementia the patient exhibits. In the model, the CDR® Dementia Staging Instrument medical test is used as the core indicator of dementia severity of the patient according to categorisation laid out by O’Bryant *et al.*^50^. Complications are child nodes to ‘*C: Suspected AD’* and ‘*Dementia severity’* as it is the progression of AD pathology that causes them.

##### Treatment

There is currently no cure for AD. The treatments prescribed are to address the symptoms which include acetylcholinesterase inhibitors and Memantine^51^. AD treatment is a child node to ‘*C: Diagnosed AD*’ as treatment is prescribed as a result of diagnosis. Other treatment nodes include use of antidepressants and antipsychotic medication, which are child nodes to ‘*CC: Depression*’ and ‘*Delusions/Hallucinations severity’*, respectively.

##### Risk Factors

The risk factor idiom models the uncertain relationship between a risk factor and the condition it affects. As such, the risk factor nodes are parents to ‘*C: Suspected AD’*. The most notable risk factors for late-onset AD are old age, ε4 allele of the apolipoprotein E gene (OPOE4) and a family history of AD. With an increase in age AD prevalence rises dramatically, estimates of 13.1% for individuals aged 75-84 years old increasing to 33.3% for individuals aged 85 and above^11^. The NIA-AA and the NICE do not recommend APOE4 genetic testing for the clinical diagnosis of AD and maintain that it should be primarily restricted to research settings^36,52^. Thus, this variable is not included in the BN. Regarding family history, a study using the Utah population database found that those who have had a first degree relative (parent or sibling) with AD are at increased risk of developing the disease compared to individuals without a first degree relative with AD^53^. It is worth considering that non-genetic factors related to family history (for example diet) may be partly responsible for the increase in risk^4^.

The concordance rate for AD in monozygotic twins is 67%^54^, indicating that a portion of the risk of AD is attributable to environmental factors. There are many reported environmental (or environmental related) risk factors including but not limited to: cardiovascular disease, smoking, diabetes, high blood pressure, traumatic brain injury (TBI), diet, and obesity^28–31^. One of the more highly recognised risk factors is TBI, thus we included it in this BN. TBI has been linked to an increased risk of AD in Danish study^59^ and in a Swedish study^60^.

Harbouring a pathogenic mutation in either the *APP, PSEN1* or *PSEN2* gene virtually guarantee the individual will develop early-onset AD^61,62^. According to the NIA-AA criteria, evidence of one of these causal mutations increases the certainty of AD^36^. Down syndrome also increases the risk of early-onset AD due to the extra copy of chromosome 21 which contains the *APP* gene^52^. There is slightly different guidance for diagnosing AD for those with learning difficulties^63^. Thus, Down syndrome is not included as a risk factor and this BN is not suitable for the diagnosis of AD for Down syndrome patients. The full BN structure is shown in Appendix B, Figure 4, but the main components are shown in Figure 3 (the difference being that, to reduce the visual complexity, the nodes ‘*Age*’, ‘*65 and above’*, ‘*C: Other dementia’* and ‘*C: Reversible condition’* are hidden in the model).

**Figure 3.**
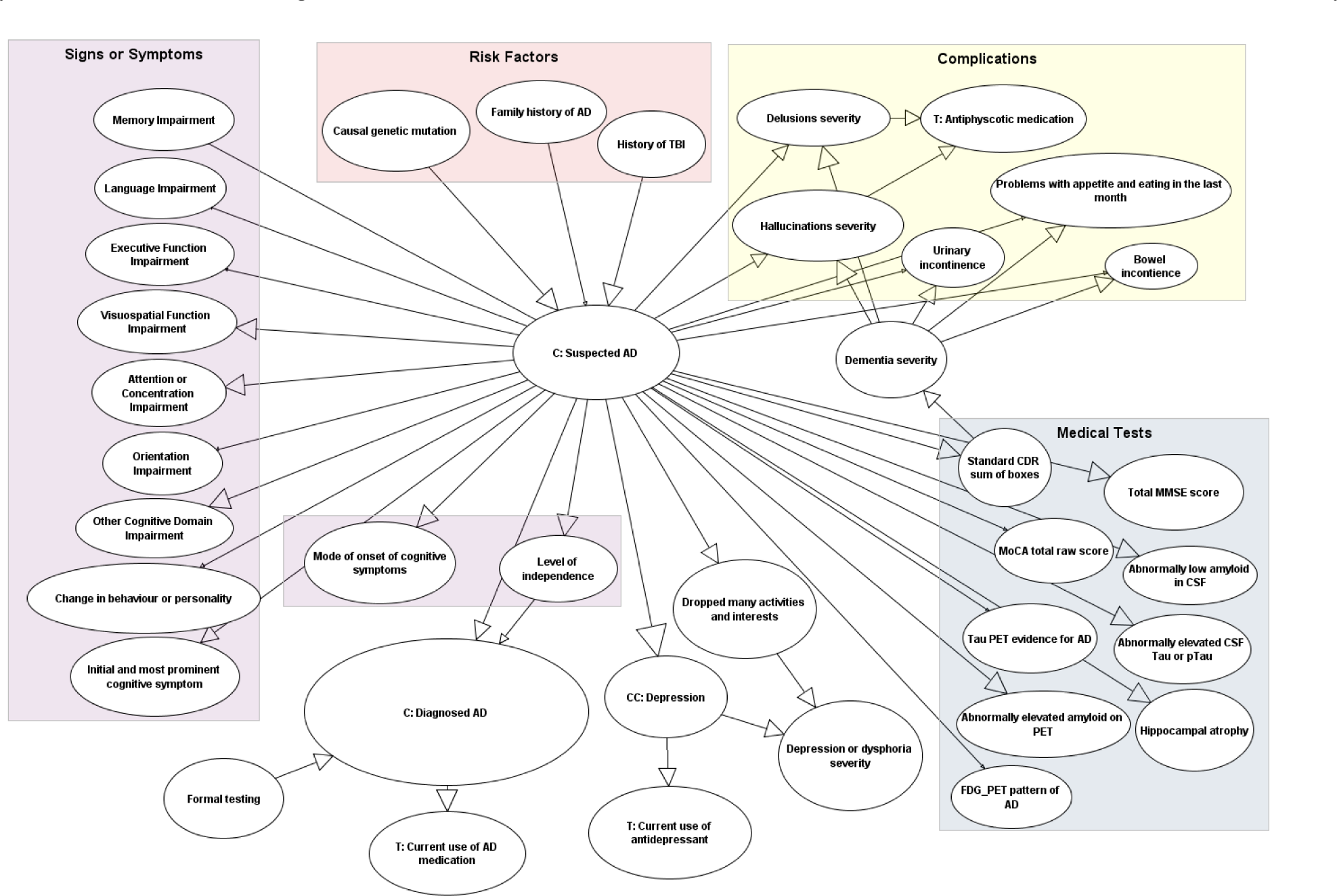
Full structure of the Bayesian network model for the diagnosis of Alzheimer’s disease. The nodes ‘*Age*’, ‘*65 and above*’, ‘*C: Other dementia*’ and ‘*C: Reversible condition’* are hidden in the model to reduce the visual complexity.

**Figure 4.**
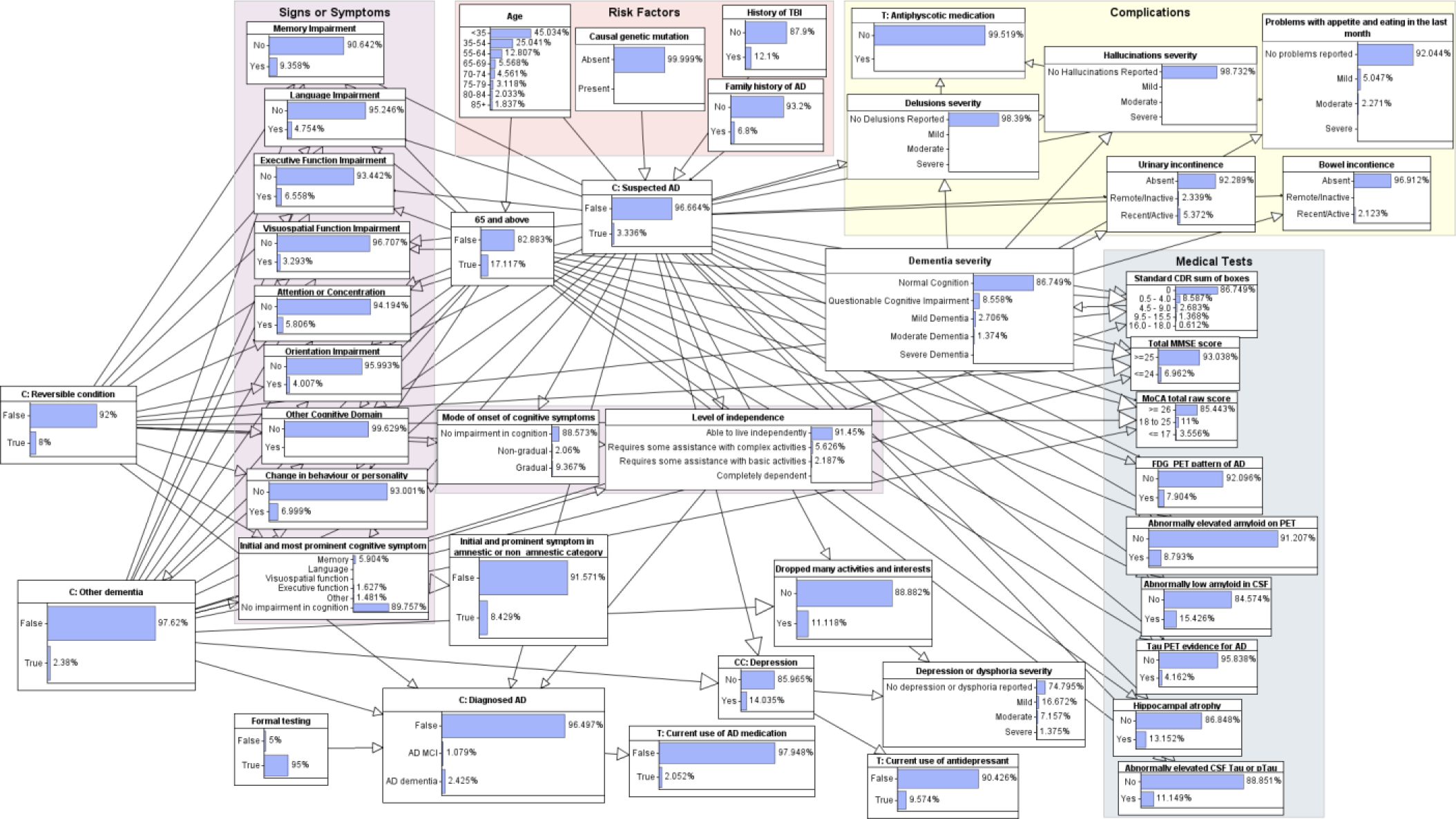
Full structure of the Bayesian network model for the diagnosis of Alzheimer’s disease.

#### 3.2.2 Specifying Parameters

##### Risk Factors

We used multiple data sources to estimate the prior probabilities for the risk factor variables. As these are root nodes, the NPTs are simply the observed marginal probability distributions. The prior probabilities for ‘*Age*’ were taken from U.S Census Bureau 2021 data^64^. The remaining parameters for the risk factor variables were extracted from the literature. The prior probability for ‘*Family history’* was obtained from a study on the Utah population database which showed that out of 270,818 residents, 6.8% had at least one first degree relative with AD^53^. The prior probability of a history of TBI was taken from a meta-analysis study including 25,134 adults which found that 12.1% had a history of TBI^65^. The prior probability of a mutation in *APP, PSEN1* or *PSEN2* was derived from considering two studies^66,67^ which, taken with other data gave a prevalence of approximately 0.000827%^66^ and 0.000201%^67^ (see Appendix A for further details). The high variation is likely because these mutations are exceedingly rare, so the prevalence rates are affected by the law of small numbers. A probability of 0.0006% was decided on but should be interpreted with caution.

##### Conditions

The NPT for ‘*C: Suspected AD’* is the conditional probability distribution for suspected AD given its risk factors. This NPT was filled in manually. The parameters were specified using judgement based on the following: those with a pathogenic mutation in *APP, PSEN1* or *PSEN2* are virtually guaranteed to develop AD and do so typically between 30-60 years of age^61,62^, the reported prevalence of AD dementia by age^11^, the prevalence of AD MCI^68^, reports of the increase in risk for those with a family history of AD^69,70^ and history of TBI^59^. The weaknesses of these reports were also considered. The parameters were calibrated to reach the prevalence of AD in the general population, 3.336%, which was calculated based on various studies (see Appendix A for further details).

For the prior probability of ‘*C: Other dementia’*, an approximate figure was derived through considering that the World Health Organisation states that AD may account for 60-70% of dementia cases^71^. We estimated that the prevalence of AD is 3.336%, assuming AD accounts for 60% of dementia cases the prevalence of other dementia is:

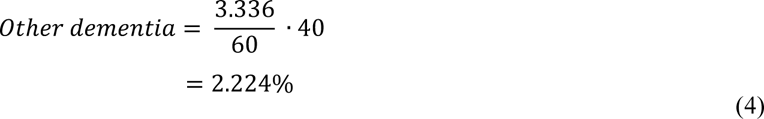

We then considered the prevalence of progressive neurological conditions that would not typically be categorised as dementia. This includes Huntington’s disease, Parkinson’s disease, and Multiple System Atrophy (MSA). Data in the literature indicate prevalence rates of 0.00271%^72^, 0.15%^73^, and 0.0019%^74^, respectively. Taking this into account, the estimated prevalence for ‘*C: Other Dementia’* (including progressive conditions not typically labelled as dementia) is 2.379%. Taking data from a 2003 study the prevalence of dementia in those under the age of 65 was found to be 0.054% of which 66% were non-AD dementia, giving an estimated prevalence of other dementia in those aged under 65 of 0.036%^75^.

Several conditions are included in the ‘*C: Reversible condition’* node including rare conditions, such as normal pressure hydrocephalus (estimated prevalence 0.01-0.02%^76^), as well as conditions such as B12 deficiency (estimated prevalence of 6% in those under 60 years of age^77^). An approximate figure of 8% was decided on for the aggregated prevalence of all the conditions included (see Appendix A Table 1).

##### Remaining Variables

The node ‘*Formal testing*’ refers to whether or not an individual has undergone the relevant examination and testing from a clinician. The probability of formal testing will vary greatly depending upon the area and many other factors. It was estimated that approximately 95% of the population would receive formal testing. The remaining variables were parametrised using the NACC UDS. This is a longitudinal dataset that includes symptom, medical test, and diagnostic data for participants of Alzheimer’s Disease Centers (ADCs) across the US. This analysis used data from 46 ADCs for UDS visits conducted between September 2005 and May 2022. Participants include individuals diagnosed with AD, individuals with other causes of cognitive impairment, and cognitively normal individuals. However, the dataset is not representative of the general population and selection biases should be considered. For example, some ADCs restrict participants to only those that agree to a post-mortem autopsy, and the cognitively normal participants tend to be highly educated volunteers^78^ who are more often aged 65 or above.

The NACC dataset was processed using Python software to select the required variables and the appropriate records. Further details on the processing steps can be found in Appendix A. The ‘Learning from Data’ tool in agena.ai was used to learn the NPTs for all remaining variables including signs/symptoms, medical tests, complications, and treatments. Due to the nature of the NACC dataset being heavily biased towards individuals with AD and other causes of cognitive impairment^78^, as well as older cognitively normal individuals, it was noted that the NPT entries learnt from data where all conditions were False and 65 and above was False were not accurate. This is because the NACC dataset is not representative of cognitively normal individuals under the age of 65. Therefore, for certain variables these NPT entries were manually entered using knowledge-based judgment, for other variables data rebalancing was performed before learning the parameters from data. NPT values learnt from data for ‘*C: Diagnosed AD’* were adjusted to reflect the possibility of undiagnosed AD and misdiagnosed AD. For further details on how the variables were parametrised see Appendix A.

## 4. Validation and Results

The key internal validation method for BNs such as this, which have been constructed using a combination of data and knowledge, is that the prior marginals of nodes with parents in the model are consistent with real world observations. What this means is that for any node X in the model, the probability distribution for X when the model is run without any observations, should be consistent with real-world observations of X. In this model the key nodes for such validation are ‘*C: Suspected AD’* and ‘*C: Diagnosed AD’*. As we only observe those diagnosed with AD we cannot formally validate ‘*C: Suspected AD’*, but it is reasonable to expect that the prior marginal will be similar to that of ‘*C: Diagnosed AD’* since undiagnosed and misdiagnosed cases are likely to be reasonably balanced.

So, the prior probability of ‘*C: Suspected AD’* and ‘*C: Diagnosed AD’* should reflect the prevalence of AD in the general population. There was difficulty in obtaining the prevalence of AD as this required obtaining rates for AD dementia specifically (instead of all-cause dementia) as well as AD MCI. Prevalence rates were taken from studies on the US population because studies on the UK population gave prevalence of dementia but not specifically AD. Moreover, there are studies on the US population that give estimates for MCI AD, which to the best of our knowledge are not available for the UK population. The prevalence of AD (including AD dementia and AD MCI) was derived from numerous studies^8,12,68,79,80^ to extract an approximate figure of 3.336% (see Appendix A for further details). As is shown in Figure 3, when no information is observed, the probability of suspected AD is 3.336%. Hence the model is internally valid.

For external validation, since the primary objective of the model is to estimate the probability of AD when certain risk factors, signs or symptoms, medical tests and other conditions are observed, we ran multiple clinical scenarios to determine whether the predictions were consistent with clinical judgements. A second type of external validation was to determine if the model could provide useful insights that are not possible with black box models. The following examples illustrate both types of validation.

### Clinical scenerio A

An individual aged 78 reports impairment in memory, attention or concentration and executive function and a change in behaviour, with no other symptoms. Entering this information in the model gives a probability of suspected AD at 67%. At this stage there is still much unknown information, observing more variables through clinical evaluation provides a more accurate estimate. Typically, when someone presents with symptoms of AD, other possible causes are ruled out before making a diagnosis^36^. If for this same individual we observe that reversible conditions are false, but another form of dementia is present, the probability of suspected AD decreases to 31% (Figure 5). This demonstrates the property of ‘explaining away’. Here the symptom nodes have several parents, if one of the parent nodes is observed as True, the probability of other parent nodes decreases, as the symptoms are most likely attributable to the parent observed as True (in this case other dementia). The clinician could then decide to conduct further biomarker tests. This would either further rule out AD, or increase the probability of dementia with mixed etiology, if negative or positive tests are observed, respectively.

**Figure 5.**
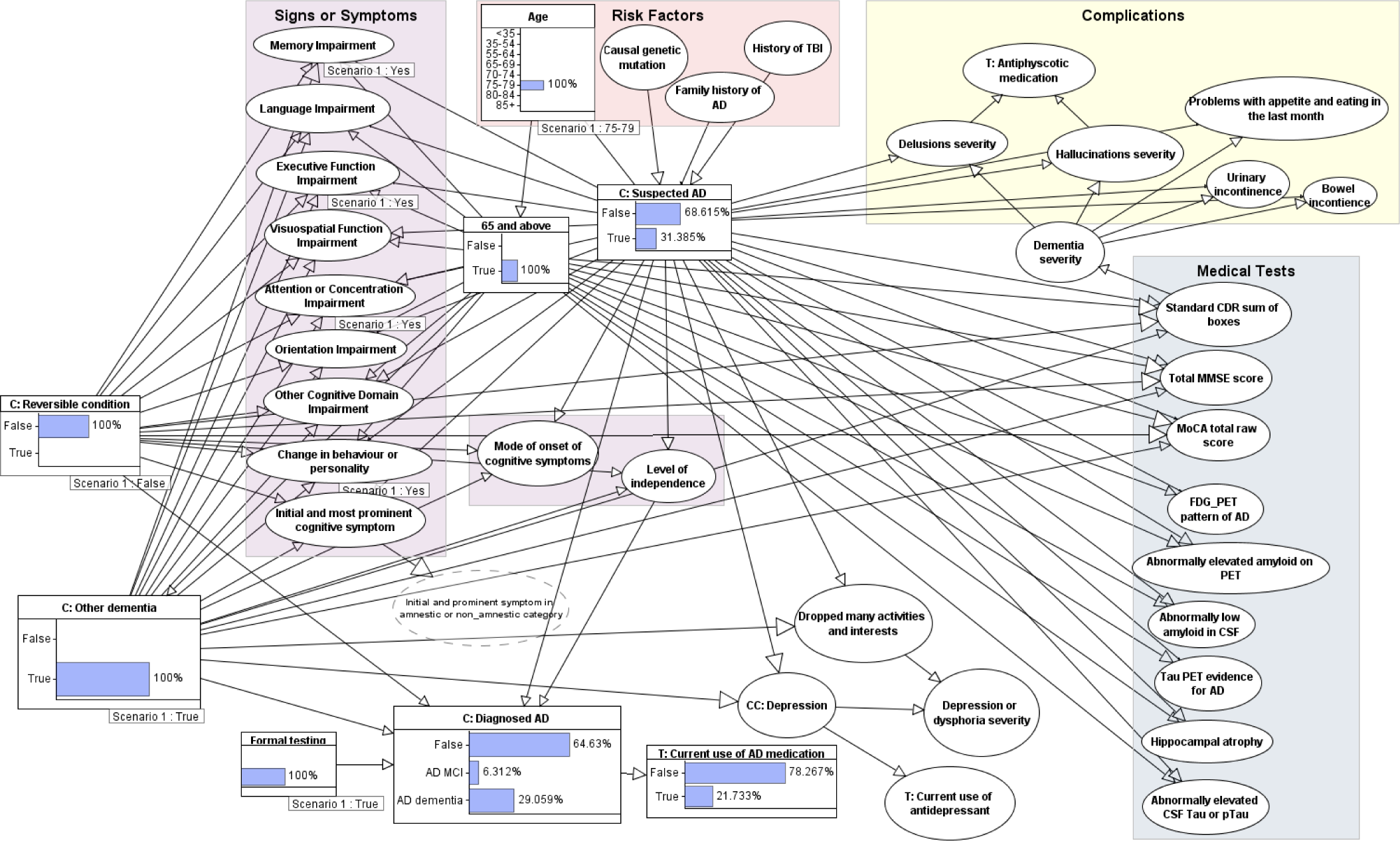
Bayesian network for clinical scenario A.

### Clinical scenerio B

An individual aged 78 presents symptoms of impairment in memory, language, and a change in behaviour, with no other symptoms. Reversible conditions are ruled out giving an initial probability of suspected AD at 65% and other dementia at 48%. A clinician may decide to conduct biomarker tests through a lumbar puncture to see whether there is pathological evidence of AD. If abnormally low amyloid in CSF and abnormally elevated CSF tau is observed (two positive biomarker tests), the probability of suspected AD is raised to 95%. The probability of AD being diagnosed is 23% for MCI and 72% for dementia, reflecting how medical tests can support a diagnosis. As the certainty of AD has increased, we again observe explaining away behaviour as the probability of other dementia reduces to 24% (Figure 6). If the level of independence is observed as ‘Able to live independently’ the probability of being diagnosed with AD MCI raises from 23% to 57%, demonstrating how the level of independence affects a diagnosis of MCI versus dementia.

**Figure 6.**
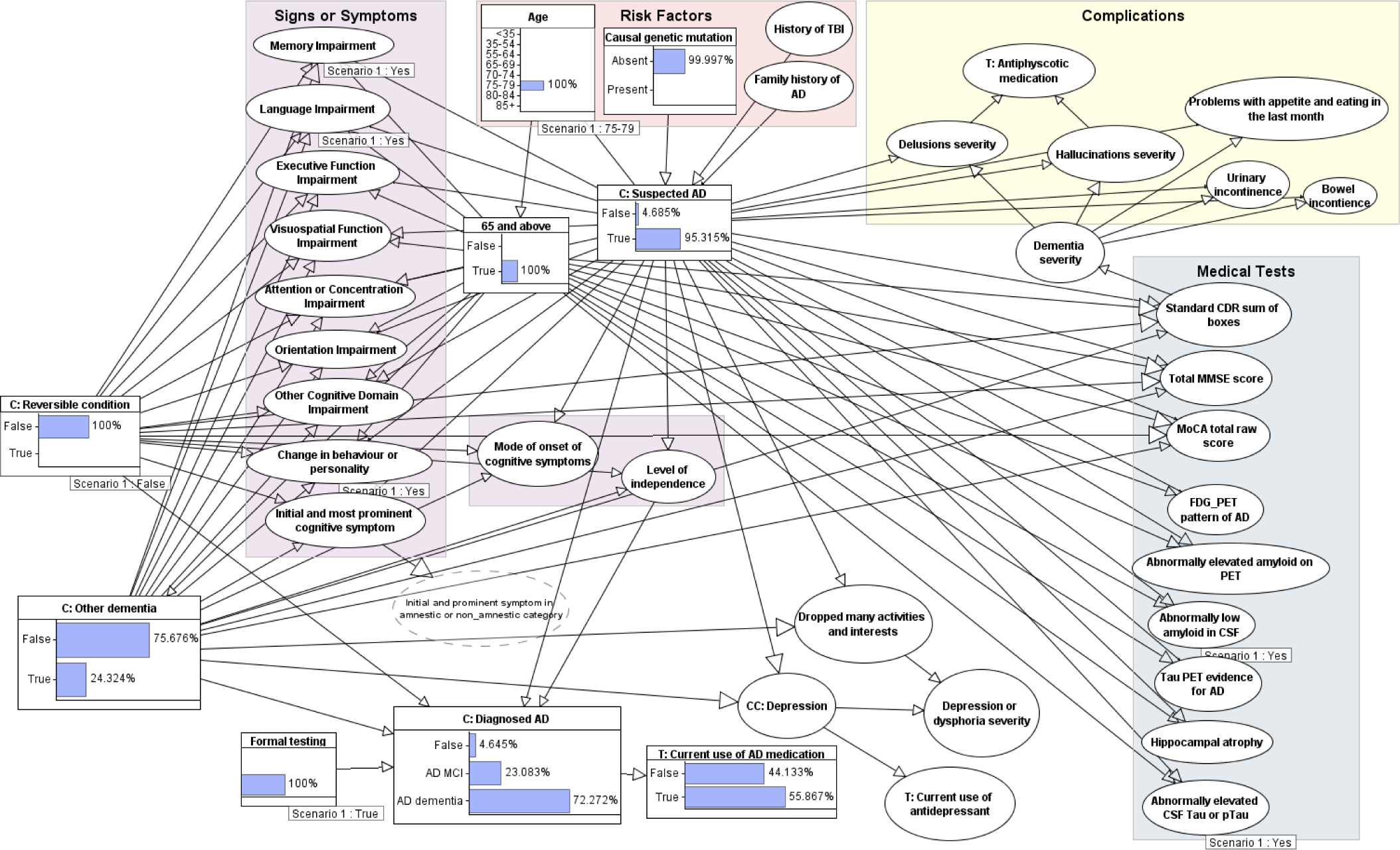
Bayesian network for clinical scenario B.

### Causal mutation and age categories under 55

Observing an age category of <35 gives a low probability of suspected AD at 0.002%, reflecting the rarity of AD in individuals below 35. Observing a causal genetic mutation raises the probability of suspected AD to 15%. When the age category is changed to 35-54 with the causal mutation still observed, the probability of suspected AD is almost certain at 99.99%. This is because these mutations cause early-onset AD which typically presents in the patients 40s or 50s, and less commonly presents in their 30s. When the only risk factor observed is an age category of <35 and signs or symptoms of impairment in memory, executive function and language are observed, and other conditions are observed as false, the probability of suspected AD is 97%, and the probability of a causal genetic mutation is increased from the marginal probability of 0.0006% to 5.139% (Figure 7). This demonstrates the ability of the model to perform backwards reasoning.

**Figure 7.**
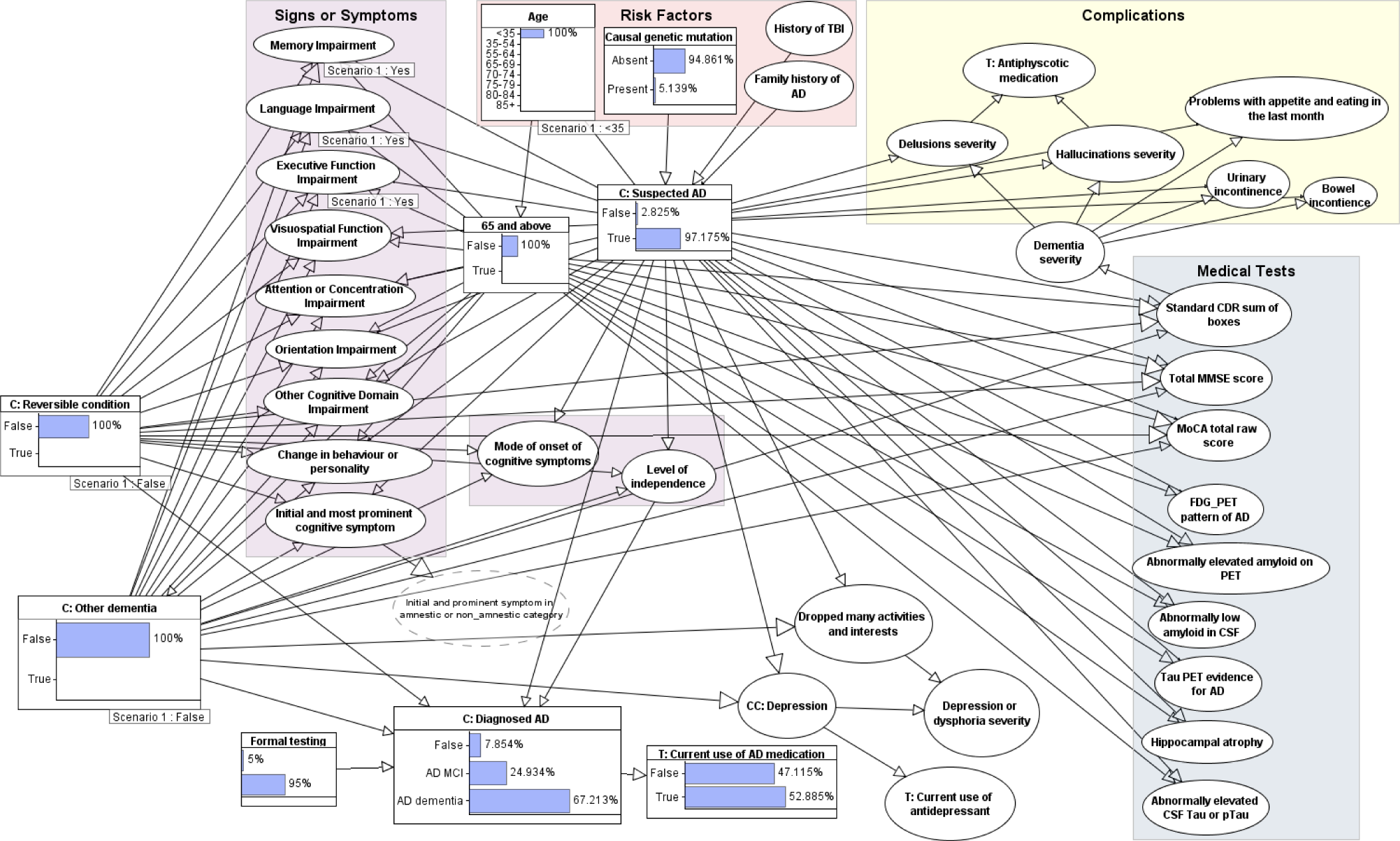
Bayesian network with age and certain signs or symptoms observed. Other conditions are ruled out (observed as False).

### Signs or symptoms and treatment

When all the signs or symptoms (except level of independence and other cognitive domain impairment) are observed, the probability of AD is 78%, and the probability of other dementia is 45%. If other dementia is observed to be false, the probability of AD is almost certain at 99.995%. The probability of each complication is greater than 8%, with the least likely complication being hallucinations and the most likely being urinary incontinence and problems with appetite and eating (individual probabilities can be seen in Figure 8). If AD diagnosis is true for MCI, the probability of treatment for AD symptoms is 27%, this raises to 69% when the diagnosis is True for AD dementia.

**Figure 8.**
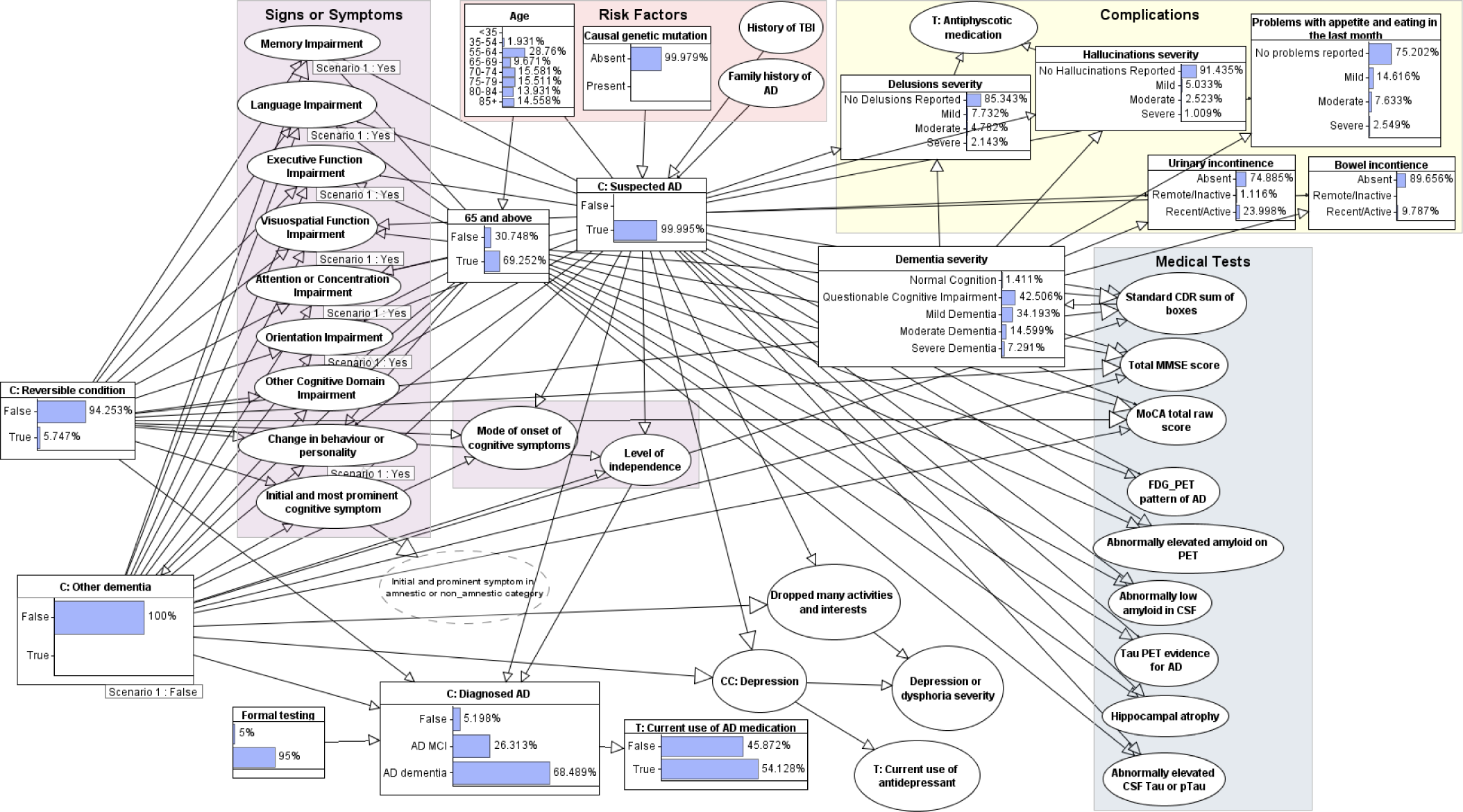
Bayesian network with all signs and symptoms observed. Other dementia is observed as False. Probabilities of complications are shown.

## 5. Conclusion, Discussion and future work

This work provides a model that can be used as a complementary tool for AD clinical diagnosis. The signs or symptoms and medical tests variables are taken from validated AD diagnostic criteria to comply with clinical practice. The model also provides the probability of the patient receiving treatment and experiencing complications. Unlike other machine-learnt (black box) AI models, this model does not require all ‘input’ variables in order to make a prediction – in any given scenario predictions are revised as more known variables are entered. Also, the model provides a visible and auditable justification for its predictions and can be used for multiple types of ‘what if analysis’. An online version of the model is available at https://ad-diagnostic-tool.public.agenaai.app/.

We estimated parameters using an approach that combines learning from data, extracting data from literature and knowledge-based judgement. There are nuances to consider in cases where a) cognitively normal individuals show the biomarker signs of AD^45^, b) those with the symptomatic profile of AD do not present the biomarker signs of AD and c) a patient exhibits dementia with mixed aetiology^36^. For these reasons, any BN proposed for the clinical diagnosis of AD should be interpreted with caution and used as a complementary tool alongside a medical professional’s evaluation of the patient.

The primary weakness of the model is the simplistic modelling of non-AD conditions that can cause cognitive impairment. These are divided into two broad categories, other dementia, and reversible conditions. Firstly, there is ambiguity for which category certain conditions belong to which could cause confusion for a prospective clinician who uses the model. For this reason, all conditions included in each category are stated in Appendix A Table 1. More prominent is the issue of aggregating multiple conditions together, which results in less accurate probabilistic modelling. An individual must only have one of the conditions belonging to the category of ‘*Other dementia*’ for its state to be set as True. For example, patient A could have Multiple-System Atrophy and patient B could have Lewy bodies dementia. Both patients would have True entered for the variable ‘*C: Other dementia*’, yet the probability of cognitive impairment associated with each condition varies greatly. It is estimated that 14% of MSA patients exhibit cognitive impairment^81,82^. Whereas for Lewy bodies, cognitive impairment is virtually guaranteed. Moreover, the symptomatic profile of different dementia causing conditions can vary. For example, a change in behaviour is more common in patients with behavioural variant FTD (bvFTD) than other dementia conditions^83^. The same applies for the different conditions included in ‘*Reversible condition*’. The impact of age-associated memory impairment^44^ has been partially accounted for using the ‘*65 and above’* node as a parent to the signs/symptoms nodes. ‘*65 and above’* is also a parent to ‘*C: Other dementia*’ to account for age as a risk factor for dementia. However, this is still simplistic in the sense that all age categories are aggregated into two broad age ranges. Moreover, it does not account for differences in age risk profile between different causes of dementia. For example, FTD can develop at a younger age to other dementia conditions^84^. Additionally, no other risk factors have been modelled for the non-AD conditions, the prevalence of these conditions vary by age^77^ and other risk factors.

Overall, there is variability between conditions in the probability of causing cognitive impairment and the symptomatic profile. Aggregating them together gives potential for statistical paradoxes to arise and ultimately results in poorer model performance. Currently, these categories are too coarse, making them more granular through modelling each condition separately would enable more accurate ‘explaining away’ behaviour and provide a more comprehensive overview of the diagnostic process. Thus, future work should seek to comprehensively model individual causes of cognitive impairment. This would include modelling risk factors and medical test idioms specific to each condition. For simplicity, the most common forms of dementia (vascular dementia, bvFTD, and Lewy Bodies dementia) should be comprehensively modelled. We will also seek to derive more accurate estimates for the prior probability of non-AD conditions through considering published estimates for each condition. Difficulty lies in that it is possible for an individual to have multiple forms of dementia and reversible conditions simultaneously.

Another weakness lies in our reliance on data exclusively from individuals diagnosed with AD to set the parameters of the child nodes for *’C: Suspected AD*.’ The core problem here is that our dataset is restricted to individuals who have received a formal AD diagnosis, excluding those who may have the disease but remain undiagnosed, such as individuals lacking access to healthcare. Consequently, while our model distinguishes between having AD and receiving a formal AD diagnosis, it does not accurately account for the rates of misdiagnoses and undiagnosed cases. Determining whether an individual received an incorrect diagnosis based on the NACC dataset is a complex task. Future research efforts could focus on determining correct and incorrect diagnoses, involving an analysis of variables related to definitive AD diagnoses (obtained through post-mortem autopsy data) and comparing them with the most recent diagnosis prior to death. It is important to acknowledge that this approach may introduce bias towards older populations. A related consideration is that while our proposed BN incorporates variables outlined in the NIA diagnostic criteria, a more robust approach would involve modeling *’C: Suspected AD’* using post-mortem autopsy data for definitive AD diagnoses. This approach avoids the limitations associated with the diagnostic criteria for possible and probable AD. By doing so, we can better use the probability of individuals experiencing specific symptoms and medical test results given they were definitively diagnosed with AD, rather than relying on those diagnosed with probable or possible AD.

Another model limitation is the lack of certain causal relationships, such as that between age and family history of AD. In our model, a family history of AD is characterised by the presence of a parent or sibling with AD. This relationship is influenced by the individual’s age: younger individuals are more likely to have parents and siblings in age categories associated with lower AD risk. There are additional variables that could be added that would improve model performance. Reports emphasise that early-onset AD is not just AD before the age of 65 and that it is often underdiagnosed and poorly managed^3^. Future work could include expanding the model to consider additional details for early-onset AD besides an age cut off and causative mutations. This includes differences in biomarker tests and symptom presentation. Another weakness is the manual setting of certain parameters based on judgment rather than data. While this approach is valid and can result in more accurate probabilistic modelling than relying solely on data, it would be more suitable to have such parameters determined by a medical professional specialising in Alzheimer’s disease diagnosis. A final noteworthy limitation is the uneven distribution of samples across the cells of the NPT, where each cell contains a conditional probability. This discrepancy becomes more apparent in nodes with multiple parents, where the number of samples satisfying each condition can vary significantly, with some having much smaller sample sizes than others.

The impact of AD and other neurological conditions on the patient, family and caregivers cannot be understated. Multiple studies have demonstrated that taking a proactive approach to managing AD and other dementias can enhance the quality of life for those affected and their caregivers^11^. Another possible area for future work involves expanding the model to assist with AD management. Here we could include variables for non-drug therapies, such as reminiscent therapy, for the management of symptoms and complications. This expansion would consider data from trials that demonstrate the efficacy of such therapies. Leveraging the causal nature of the model, we would be able to model interventions. For instance, we could ask questions such as: If a patient is prescribed a specific medical treatment, what is the probability they will experience symptoms or complications?

It should be noted that some sites state the definitive diagnosis of AD still requires autopsy, while others say it is possible with biomarker testing. Meanwhile the nomenclature surrounding AD has driven revised definitions. Later work in 2018 by the NIA-AA posits a purely biological definition of AD which is only to be used in research settings^85^, and ^86^ highlights the distinction between AD as a disease and AD as a clinical syndrome. Heterogeneity within the literature exists, though at the core there is an emphasis of the distinction between the clinical diagnosis of AD and the diagnosis of AD for research purposes. This is driven in part due to the rise in biomarker testing and evidence suggesting they may be effective in predicting pre-clinical AD, a stage of AD with significant research interest for prospective treatments. In this case there is a stronger emphasis on the underlying biology of the disease for the treatment to target a particular pathophysiological process. Future work could include developing an AD diagnostic BN tool for research purposes that includes preclinical AD. This BN would include genetic risk factors (such as OPOE4), mitigants (such as OPOE2 and others)^56,87^, and a stronger weight for biomarker evidence.

## Data Availability

Data obtained from the National Alzheimer's Coordinating Center are available upon request online at https://naccdata.org/

## Acknowledgment

We are thankful to the NACC for providing us with the NACC dataset.

The NACC database is funded by NIA/NIH Grant U24 AG072122. NACC data are contributed by the NIA-funded ADCs: P50 AG005131 (PI James Brewer, MD, PhD), P50 AG005133 (PI Oscar Lopez, MD), P50 AG005134 (PI Bradley Hyman, MD, PhD), P50 AG005136 (PI Thomas Grabowski, MD), P50 AG005138 (PI Mary Sano, PhD), P50 AG005142 (PI Helena Chui, MD), P50 AG005146 (PI Marilyn Albert, PhD), P50 AG005681 (PI John Morris, MD), P30 AG008017 (PI Jeffrey Kaye, MD), P30 AG008051 (PI Thomas Wisniewski, MD), P50 AG008702 (PI Scott Small, MD), P30 AG010124 (PI John Trojanowski, MD, PhD), P30 AG010129 (PI Charles DeCarli, MD), P30 AG010133 (PI Andrew Saykin, PsyD), P30 AG010161 (PI David Bennett, MD), P30 AG012300 (PI Roger Rosenberg, MD), P30 AG013846 (PI Neil Kowall, MD), P30 AG013854 (PI Robert Vassar, PhD), P50 AG016573 (PI Frank LaFerla, PhD), P50 AG016574 (PI Ronald Petersen, MD, PhD), P30 AG019610 (PI Eric Reiman, MD), P50 AG023501 (PI Bruce Miller, MD), P50 AG025688 (PI Allan Levey, MD, PhD), P30 AG028383 (PI Linda Van Eldik, PhD), P50 AG033514 (PI Sanjay Asthana, MD, FRCP), P30 AG035982 (PI Russell Swerdlow, MD), P50 AG047266 (PI Todd Golde, MD, PhD), P50 AG047270 (PI Stephen Strittmatter, MD, PhD), P50 AG047366 (PI Victor Henderson, MD, MS), P30 AG049638 (PI Suzanne Craft, PhD), P30 AG053760 (PI Henry Paulson, MD, PhD), P30 AG066546 (PI Sudha Seshadri, MD), P20 AG068024 (PI Erik Roberson, MD, PhD), P20 AG068053 (PI Marwan Sabbagh, MD), P20 AG068077 (PI Gary Rosenberg, MD), P20 AG068082 (PI Angela Jefferson, PhD), P30 AG072958 (PI Heather Whitson, MD), P30 AG072959 (PI James Leverenz, MD).

## Appendices

### Appendix A. Deriving the prevalence of AD and causative mutations, NACC data processing, BN variables

In this section, we outline the methodology used to derive prevalence estimates for Alzheimer’s Disease (AD) and the prevalence rates of causative mutations related to Autosomal-dominant early-onset AD (ADEOAD). The prevalence estimates are derived based on available data.

### AD prevalence estimation

For these calculations the following numbers were taken from the US census bureau 2021 data:

- Total number of people aged 35-64 = 123,459,000
- Total number of people aged 65 and above = 55,836,000
- The total US population = 326,195,000

To estimate the prevalence of AD in the United States, we employed a data-driven approach using figures from the US Census Bureau’s 2021 data as well as prevalence studies. We considered two categories within AD: AD Mild Cognitive Impairment (MCI) and AD Dementia.

#### AD MCI

According to the Health and Retirement Study (HRS) that classified participants based on clinical symptoms, approximately 22% of individuals aged 65 and above have MCI^88^. Biomarker and PET scan studies suggest that around 50% of individuals with MCI exhibit AD-related brain changes^68,80^. By combining these findings, we estimated that 11% of individuals aged 65 and over have AD MCI. This corresponds to 1.883% prevalence within the entire US population. We acknowledged the potential underrepresentation of adults under 65 with AD MCI and consequently used a slightly higher estimate.

#### AD Dementia

Projections from a population study based on data from the Chicago Health and Aging Project gave an estimate of 6.7 million individuals with AD dementia in the US^8^. However, numerous biomarker studies from both autopsies and clinical trials indicate that individuals classified as having AD based on symptoms do not always present biology brain changes characteristic of AD (between 15-30%)^11^. Adjusting for this discrepancy, we estimated that 4.7 million individuals have AD dementia, resulting in a 1.441% prevalence rate.

#### AD Dementia under 65 (EOAD)

Both the studies on AD MCI and AD dementia consider adults aged 65 or over. For the EOAD prevalence estimate, we incorporated the population aged 35-64. Using a meta-analysis, we found a prevalence rate of 31.8 per 100,000 for EOAD in this age group^79^, projecting to 39,260 affected individuals in the US. This gives a prevalence of 0.012% when considering the total US population. A limitation is that those aged below 35 were not considered in this estimate.

Considering the prevalence rates for AD MCI and AD dementia in individuals aged 65 and over, as well as EOAD, we calculated an overall AD prevalence of 3.336%. It is important to acknowledge that while we used US data for these calculations, the generalisation to the entire population is a limitation, as AD prevalence rates vary among different countries.

### APP, PSEN1 and PSEN2 mutation prevalence estimation

Based on Campion *et al*.^62^, we calculated a prevalence of ADEOAD at 0.00117%. This figure was derived from identifying 5 out of 426,710 residents in Rouen who met the criteria for ADEOAD, defined as the occurrence of AD cases with onset at age <61 years in three generations^62^. In the same study, mutational analysis of *APP, PSEN1*, and *PSEN2* was performed in 34 families with ADEOAD, including the 5 initially identified and additional referred families. Among these 34 families, 24 out of 34 (70.6%) had mutations in *APP* or *PSEN1*. When calculating 70.6% of 0.00117% without rounding, this equals 0.0008271%.

From Jarmolowicz *et al*.^67^, we determined a prevalence of ADEOAD to be 0.00172%. This estimation was based on their observation that 17 out of 120 (14.2%) Australian EOAD patients had ADOEAD, in conjunction with a previously established prevalence of EOAD at 0.012% (Hendriks *et al*.)^79^. Calculating precisely, 14.2% of 0.012% gives a prevalence of ADEOAD at 0.0017%. In the same study, mutational analysis of the 17 individuals with ADEOAD revealed that 2 out of 17 (11.8%) had known mutations. Using the non-rounded figures for calculations, this results in 0.000201%.

### NACC data processing

The unprocessed NACC dataset has over 750 variables and one row of data per subject visit (169,408 rows), meaning a sole participant can have multiple rows of data. The first step was to narrow down the variables to those required for the BN. This involved a combination of selecting the relevant variables and deriving a single variable from multiple variables. When deriving a single variable from multiple variables, variables with a large amount of missing data were considered. For example, ‘*C: Other dementia’* was derived from 21 variables, a large proportion of the data had missing values for cerebrovascular disease, vascular dementia, posterior cortical atrophy, MSA and FTLD. Providing that none of the ‘Other dementia’ conditions were True, if a subject was False for the 7 other conditions included in ‘Other dementia’ and the data was missing for any or all of the 5 aforementioned conditions, the subject was labelled as False for ‘Other dementia’. Next, the samples were decreased by only keeping the sample for each participant with the least missing values. This ensured there were no duplicate subjects. A total of 37 variables were used from the NACC dataset to parametrise the BN, data subjects with more than 18 (over 50%) missing values were dropped. The resulting dataset has 45,776 samples, 18,1807 (41%) of which are classified as having AD, and 28740 (63%) are classified as having either AD, other dementia, or a reversible condition.

For learning from data using the NACC dataset, missing values were estimated using an EM algorithm. Except where variables had more than 5,000 missing samples. In this case conditional probabilities were calculated after dropping samples with missing data for each individual variable. The following variables had over 5,000 missing samples: all biomarker tests, MMSE, MoCA and Orientation impairment.

For biomarker tests, it was non-trivial to find the probability of a positive biomarker test given AD = False and 65 and above = False. As a result, the NACC dataset was used to learn these values. We discussed previously that the NACC dataset is not representative of the population under these criteria, primarily because they are more likely to have other causes of cognitive impairment. To address this, we first created datasets for each biomarker test where entries with missing test results for a given test were dropped. Considering that some of the ‘Other dementia’ conditions can affect certain biomarker tests, and that the estimated prevalence of other dementia under 65 is 0.036%, we then dropped entries where ‘*Other dementia’* = True iteratively until a percentage of 0.05% of subjects having ‘*Other dementia’* = True was reached (note that due to the smaller proportion of samples, there was often no entries where ‘*Other dementia’* = True). We then recalculated the probability of a positive biomarker test given AD = False and 65 and above = False which were populated in the respective NPT fields. The sample size for each biomarker test after these processing steps is as follows: CSF tau: n=167, CSF amyloid: n=168, PET amyloid: n=203, PET tau: n=105, FDG: n =130, Hippocampal atrophy: n=1345.

### BN variables

To reduce the visual complexity, it was necessary to simplify the names of certain variables in the BN. The table below gives a description of such variables.

The conditions included in ‘Other dementia’ are categorised as such because they are potential causes of dementia, however for some it is possible that dementia is not present. We acknowledge that including MSA in the ‘Other dementia’ category when dementia is currently an exclusionary factor for diagnosing MSA^89^ may seem erroneous. However, it is included in this category because a) MSA is reported to cause cognitive impairment in approximately 14% of patients^82^ and continuing evidence that cognitive impairment is an integral part of the disease^90^ and b) MSA is not a reversible condition. We are aware of the limitations of having two simplistic categories, and in future work differentiating individual conditions will avoid this confusion. Similarly, there is cross over between cerebrovascular disease and vascular dementia, and future work will aim to make individual conditions explicitly modelled.

Although the conditions included in ‘Reversible condition’ are potential causes of cognitive impairment, cognitive impairment is not always present. It should be noted that there are other reversible conditions that can account for cognitive impairment not included in this list, for example depression. Including depression poses issues as it is a common comorbidity observed with AD. In future work we can expand to correctly model depression as both a potential cause of cognitive impairment and as a comorbidity of AD. It should be noted that in some literature certain reversible conditions may also be referred to as dementia conditions. For example, normal pressure hydrocephalus, which is often a reversible condition, is sometimes referred to as a cause of dementia^91^.

